# A Human Gait Circuit Derived from Brain Lesions and Deep Brain Stimulation

**DOI:** 10.1101/2025.03.28.25324852

**Authors:** Lan Luo, Frederic L. W. V. J. Schaper, Michael Nguyen, Sandrine Jabbour, Joey Hsu, Louis Soussand, Christopher Lin, Shan H. Siddiqi, Konstantin Butenko, Andreas Horn, Martin Reich, Jens Volkmann, Andrea A. Kühn, Maurizio Corbetta, Ron L. Alterman, Michael D. Fox

## Abstract

**Objectives:** The relevant neuroanatomy for gait dysfunction remains unclear. We sought 1) to identify a brain circuit for gait impairment post stroke, 2) to identify a brain circuit for gait changes after subthalamic DBS for Parkinson’s disease, 3) to test for convergence between these two circuits.

**Methods:** This cross-sectional retrospective study used data from four independent datasets including 109 individuals with stroke and 125 patients with PD who received subthalamic DBS. Gait impairment post stroke was measured using the Combined Walking Index. Gait changes after subthalamic DBS was measured using the gait subscore on the UPDRS Part III. Connectivity between lesion locations or DBS sites and two *a priori* locomotor regions was computed using a large normative connectome. We repeated this analysis in a data-driven fashion to identify additional connections.

**Results:** Connectivity between lesion locations and *a priori* regions in the pedunculopontine nucleus and cerebellar locomotor region were associated with gait impairment post stroke (*p*< 0.05), along with connectivity to a distributed circuit of other brain regions. Connectivity between DBS sites and our *a priori* gait regions and connectivity between DBS sites and our lesion-based gait circuit were all associated with gait changes post DBS (*p* < 10^-3^). Data-driven gait circuits derived from lesions and DBS showed similar topography and converged on a common brain circuit (spatial r = 0.68, *p* = 0.0063, 10000 permutations).

**Interpretation:** Lesion locations impairing gait and DBS sites modulating gait converge on a common brain circuit, which may provide a neuromodulation target for gait dysfunction.

## Introduction

Gait impairment is a common symptom found in a wide range of neurological disorders. For example, gait abnormalities are found in more than half of stroke survivors^1,2^ and are an important predictor of nursing home placement after hospitalization for stroke.^3,4^ However, the neuroanatomical substrate of post-stroke gait impairment remains unclear, in part because lesions in different brain locations can lead to gait impairment.^5^ This has led to the hypothesis that lesions causing gait impairment might localize to a specific brain circuit,^6,7^ but this hypothesis remains to be directly tested.

Over 80% of patients with Parkinson’s disease (PD) also have gait impairment, leading to falls and a decline in overall quality of life.^4,8–10^ Treatments that work well for other symptoms of PD such as deep brain stimulation (DBS) often fail to improve gait and can sometimes worsen gait in some patients.^11–19^ As in stroke, the brain regions or circuits involved in parkinsonian gait remain unclear, and alternative DBS targets aiming at gait improvement have led to mixed results.^20^

Gait dysfunction differs significantly between stroke and PD, including their etiology, pathology, phenomenology, and spatiotemporal signatures.^21^ However, there is reason to think that they may share a common neuroanatomical substrate. For example, regions such as the pedunculopontine nucleus (PPN) and cerebellar locomotor region (CLR) have been implicated in both post-stroke gait decline^22,23^ and DBS-induced gait improvement.^24^ Furthermore, recent investigations into other symptoms of brain disease have found that lesions that cause a particular symptom and DBS electrodes that improve that symptom can be connected to a common brain circuit. This premise has held true for parkinsonism,^25^ dystonia,^26,27^ depression,^28^ cognitive decline,^29^ tics,^30^ and epilepsy.^31^ Finding convergence across two independent causal sources of information (lesions and DBS) and across very different patient populations (stroke and PD) can increase confidence in localization of a symptom beyond what is possible with a single modality or patient group alone^32^.

In the present study, we devised three experiments using stroke lesions and DBS stimulation sites **(Figure 1A)**. First, we used two *a priori* locomotor regions, the PPN and CLR, to determine whether stroke lesion locations and DBS electrodes that modulate gait are connected to these two regions of interest (**Figure 1B**). Second, we mapped lesions causing gait impairment onto a brain circuit using a technique termed lesion network mapping (**Figure 1C**).^25,26,33–42^ This technique uses a large normative functional connectome to localize lesions onto a distributed brain circuit. Third, we mapped DBS electrodes causing gait improvement onto a brain circuit using DBS network mapping (**Figure 1C**).^25,26,28–30^ Since each patient is stimulated at a slighted different location, we can identify connections associated with an outcome of interest. Finally, we test whether these two circuits converge on a common substrate (**Figure 1D**).

**Figure 1.**
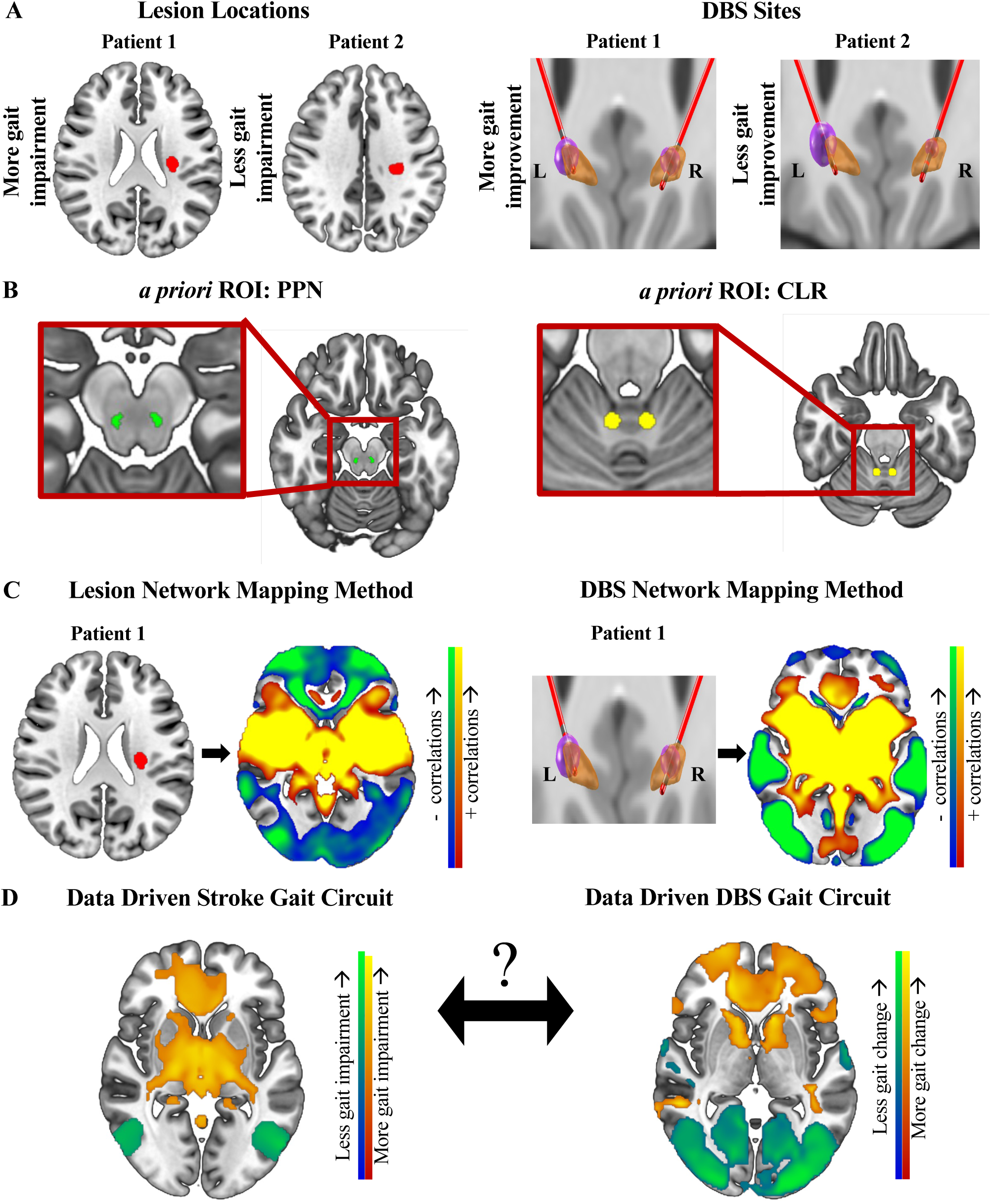
Methodology. A, B. First, we determined whether stroke lesions impairing gait and DBS sites modulating gait are connected to our *a priori* locomotor regions, the PPN and CLR. C. Using lesion network mapping, we derived a brain circuit for post-stroke gait impairment using resting-state functional connectivity data from 1000 healthy participants. Similarly, we used DBS network mapping to derive a brain circuit for post-DBS gait change. D. Finally, we determined whether these two gait circuits show convergence. PPN = pedunculopontine nucleus, CLR = cerebellar locomotor region.

## Methods

This multicenter study was carried out in accordance with the Declaration of Helsinki, approved by the institutional review board of Beth Israel Deaconess Medical Center (BIDMC), Boston, Massachusetts. Strengthening the Reporting of Observational Studies in Epidemiology (STROBE) reporting guidelines for cross-sectional studies were followed.

### Description of Lesion Dataset

We utilized a dataset consisting of 109 subjects with first time ischemic or hemorrhagic stroke who were admitted to Barnes-Jewish Hospital (St. Louis, MO) between 2008-2013. Inclusion and exclusion criteria were previously described by Corbetta *et al.*^43^ National Institutes of Health Stroke Scale (NIHSS)^44^ scores and gait scores were collected two weeks post stroke, along with a variety of other phenotypic information.^43^ The combined walking index, a measure of gait independence and speed, was used as a continuous variable throughout our analyses.^43,45,46^ Participants who were assigned gait scores one through six required assistance to walk, while those with scores seven through nine walked independently and were further characterized based on gait speed.^43,45,46^ In all our lesion analyses, we negated the combined walking index so that higher values aligned with more unfavorable gait outcomes, thus establishing consistency with the directionality of NIHSS scores. Lesions were manually segmented based on magnetic resonance imaging images, spatially normalized to Montreal Neurological Institute (MNI) 152 atlas space and binarized such that voxels within the lesion carried a value of 1 while all other voxels carried a value of 0. Supplementary analyses leveraged other phenotypic information including factor scores for motoricity, attention, right and left motor function, spatial memory, verbal memory, and language, as detailed previously.^43^

### Connectivity to A Priori Locomotor Regions

To determine if gait changes post stroke are associated with functional connectivity to two *a priori* locomotor ROIs, we tested whether functional connectivity of the 109 stroke lesions to either the PPN or CLR was associated with post stroke gait impairment. First, we generated 3-mm spherical seeds centered around PPN coordinates from Alho *et al.* and CLR coordinates based on averaging seven CLR studies as reported by Boyne *et al*.^47,48^ Next, using a publicly available normative connectome consisting of resting-state functional connectivity MRI data from 1000 healthy volunteers,^49^ we determined the functional connectivity of each stroke lesion to our *a priori* ROIs. Functional connectivity values were transformed to a normal distribution using Fisher’s r to z transform. Finally, the correlation between connectivity and post stroke gait impairment was analyzed using Pearson’s correlation coefficient with correction for stroke severity using NIHSS scores.

### Lesion Network Mapping

We used lesion network mapping, as described in our prior publications,^25,26,34–41,50,51^ to determine whether stroke lesions associated with gait impairment mapped to a common brain circuit. In short, the functional connectivity between each lesion location and all other brain voxels was computed using the same normative connectome as our *a priori* ROI analysis. Next, we compared unthresholded lesion circuit maps for individuals with varying degrees of gait impairment using a general linear model and permutation testing^52,53^ while correcting for stroke severity. Cluster-based family-wise error (FWE)-corrected *p* < 0.05 was considered significant. We will refer to this statistical map as our “***stroke gait circuit***”. Supplementary analyses tested whether the topography of this stroke gait circuit was robust to controlling for other stroke deficits (motoricity, attention, right and left motor function, spatial memory, verbal memory, and language) and whether data-driven circuit maps for any of these deficits match the topography of the stroke gait circuit.

### Controlling for Lesion Location

To test whether the above results were due to connectivity or could also been identified using lesion locations alone, we performed voxel-based lesion symptom mapping^54^ in NiiStat (version 1.1; https://github.com/neurolabusc/NiiStat)^58^ running on MATLAB version R2021a and SPM version 12. Lesioned voxels were treated as the independent variable and the negated combined walking index as the dependent variable. One-tailed statistical tests were applied based on the assumption that lesions will lead to gait impairment and not improvement. For all tests, we used *p* < 0.05 corrected for multiple comparisons with permutation thresholding (2000 permutations). We then performed adjusted lesion symptom mapping using Freedman-Lane multivariable regression approach to model negated combined walking index^52^ while correcting for lesion volume and stroke severity.

### Description of DBS Datasets

We utilized three independent bilateral subthalamic nucleus (STN) DBS datasets: 1) a previously published cohort of 51 participants with PD who underwent surgery at Charité Universitätsmedizin in Berlin, Germany,^55^ 2) another previously published group of 44 subjects who had surgery at Würzburg University Hospital in Würzburg, Germany,^55^ 3) and an unpublished cohort of 31 participants who had surgery at BIDMC in Boston, MA. Demographics and clinical information of the two German cohorts are described in a previous publication^55^ and those for the Boston cohort can be found in **Table 1**. All three DBS studies were approved by their respective Institutional Review Boards. For all participants, Unified Parkinson Disease Rating Scale (UPDRS)^56^ part III or the International Parkinson and Movement Disorder Society-sponsored revision of the Unified Parkinson’s Disease Rating Scale (MDS-UPDRS)^57^ part III scores along with gait subscores (UPDRS item 3.29 or MDS-UPDRS item 3.10) were collected for DBS *off* and DBS *on* conditions. For simplicity, we will refer to the motor sections of both versions of the UPDRS as “UPDRS-III” throughout this article. For the Boston cohort, DBS response was assessed as the change between the DBS *on* state 1-12 months postop versus the DBS *off* state during the initial programming visit, with both visits performed either after participants withdrew from dopaminergic medication for at least 12 hours (medication *off* state) or 1 hour after taking their medications (medication *on* state). Motor scores for two of the DBS cohorts (Berlin and Würzburg) were obtained in the medication *off* state. One individual from the Würzburg cohort was excluded from our analysis due to missing data (Würzburg n = 43; total n = 125).

For supplementary analyses, we also computed average scores for lower extremity bradykinesia (UPDRS item 3.26; MDS-UPDRS items 3.7a, 3.7b, 3.8a, 3.8b), lower extremity rigidity (UPDRS item 3.22; MDS-UPDRS items 3.3d, 3.3e), and postural instability (UPDRS item 3.30; MDS-UPDRS item 3.12) as described in prior work.^57^

### DBS Electrode Localization and VTA Generation

All DBS electrodes were localized using Lead-DBS software version 2.1.5 (www.lead-dbs.org),^58,59^ including brain shift correction and normalization into standard stereotactic (MNI) space using Advanced Normalization Tools (http://stnava.github.io/ANTs/).^60^ After taking into account lead localization and clinical stimulation parameters, electric fields (E-fields) were calculated using the same software as previously detailed by Horn and colleagues.^58^ These fields were thresholded at the default value of 0.2 V/m, to calculate binary volumes of tissue activated (VTAs).

### Connectivity to A Priori Locomotor Regions

Connectivity between DBS sites and our *a priori* ROIs was computed using our combined DBS dataset. Correlation between these connectivity values and the changes in gait after DBS was computed, uncorrected and corrected for dataset and change in total motor score using UPDRS-III.

### DBS Network Mapping

Using the same normative connectome as above, we first determined functional connectivity estimates between each VTA and all other brain voxels. We then identified the connectivity of VTAs independently associated with DBS gait changes by comparing unthresholded DBS circuit maps and changes in gait post DBS using a general linear model and permutation testing^52,53^ with dataset and change in total motor score as covariates.^52,53^ Cluster-based FWE-corrected *p* < 0.05 were considered significant. This analysis was first performed using each DBS cohort and then repeated using all three DBS groups combined. We will refer to this statistical map (using all three DBS cohorts) as our “***DBS gait circuit***”. We compared the spatial topography amongst all three DBS-derived gait circuits using the mean spatial cross-correlation with 10000 permutations and an alpha level of *p* < 0.05 as described previously by Siddiqi *et al*.^61^

Supplementary analyses tested whether the topography of this DBS gait circuit was robust to controlling for other DBS changes (lower extremity bradykinesia, lower extremity rigidity, and postural instability), and whether data-driven circuit maps for any of these changes match the topography of the DBS gait circuit. These supplementary analyses were conducted on a subset of patients (n = 94 of 125) for which these sub-scores were readily available (Berlin and Würzburg datasets).

### Controlling for DBS Site

To test whether the above results were due to connectivity or could also be identified based on stimulation site alone, we compared VTA locations with gait change as a binary variable (either improvement or no improvement) using a general linear model and permutation testing (Permutation Analysis of Linear Models in FSL version 6.0),^52^ correcting for both VTA size and total UPDRS-III change (defined as the difference between total UPDRS-III scores in DBS *on* versus DBS *off* states). Cluster-based FWE corrected *p* < 0.05 was considered significant. To confirm results using an entirely different approach, we utilized the DBS Sweetspot Explorer toolbox in Lead-DBS software version 2.1.5 (www.lead-dbs.org)^58,59^. Voxels were considered if at least 20% of E-fields (N=23) reached them at a magnitude 0.2 V/mm. DBS gait change, defined as the difference between DBS *off* versus DBS *on* gait scores, was used as the variable of interest and the change in total motor score was used as a covariate. Wilcoxon signed-rank testing was performed with the average post DBS gait outcome as the null hypothesis and correcting for VTA size. Voxels with *p* < 0.05 with Bonferroni correction were considered significant. This analysis was repeated with four sweetspot mapping approaches: 1) Butson *et al.* using a VTA-based mean-effect image thresholded to select voxels with an average gait improvement greater than 50%,^62^ 2) Cheung *et al.* using at least 75% of responder VTAs overlap,^63^ 3) Reich *et al.* selecting voxels with *p* < 0.05 after voxel-wise two-sample t-test and with spatial clusters of at least 500 voxels,^64^ and 4) Dembek *et al.* selecting voxels with *p* <0.05 after voxel-wise Wilcoxon signed-rank tests and with non-parametric permutation to account for multiple comparisons.^65^

### Convergence between Lesion and DBS Gait Circuits

Thus far, we have evaluated the lesion and DBS associations with gait separately. To test whether these two independent sources of information converge onto common neuroanatomy, we performed three analyses. First, we tested whether connectivity between our stroke gait circuit and STN DBS sites was associated with gait changes after DBS. Connectivity between each pair of VTAs and our unthresholded stroke gait circuit were calculated and correlated with post DBS gait changes. Correlation between the connectivity and gait outcomes was performed using Pearson’s correlation coefficient with significance of *p* < 0.05. A partial correlation was also carried out to correct for dataset and the change in total motor score. Second, we compared the spatial topography of our unthresholded stroke gait circuit and DBS gait circuit using spatial correlation. To determine if this spatial correlation was stronger than expected by chance, we re-computed the spatial cross-correlation after randomly swapping lesion locations with post-stroke gait changes and DBS sites with post DBS gait changes, using 10000 permutations at an alpha level of *p* < 0.05.^61^ Third, we tested whether any brain regions were significantly associated with gait both in the lesion analysis and the DBS analysis by performing a conjunction analysis.

Specifically, we identified voxels that were significant in both our stroke gait circuit and DBS gait circuit, which we will refer to as our “conjunction map.”

## Results

### Lesions and Gait Impairment

Two weeks post stroke, 71 lesions (65.1%) were associated with gait impairment. Lesions associated with gait impairment occurred in heterogeneous locations throughout the brain. Stroke severity, as defined by NIHSS score, accounted for 71% of the variance in gait impairment (**Supplementary** Figure 1).

### Connectivity to A Priori Locomotor Generators

Connectivity between stroke lesions and our two *a priori* ROIs was significantly associated with gait impairment (PPN: r = 0.22, *p* = 0.045; CLR: r = 0.21, *p* = 0.031), both of which remained significant after correcting for stroke severity (PPN: r = 0.22, *p* = 0.025; CLR: r = 0.23, *p* = 0.018) (**Figure 2 A-B**).

**Figure 2.**
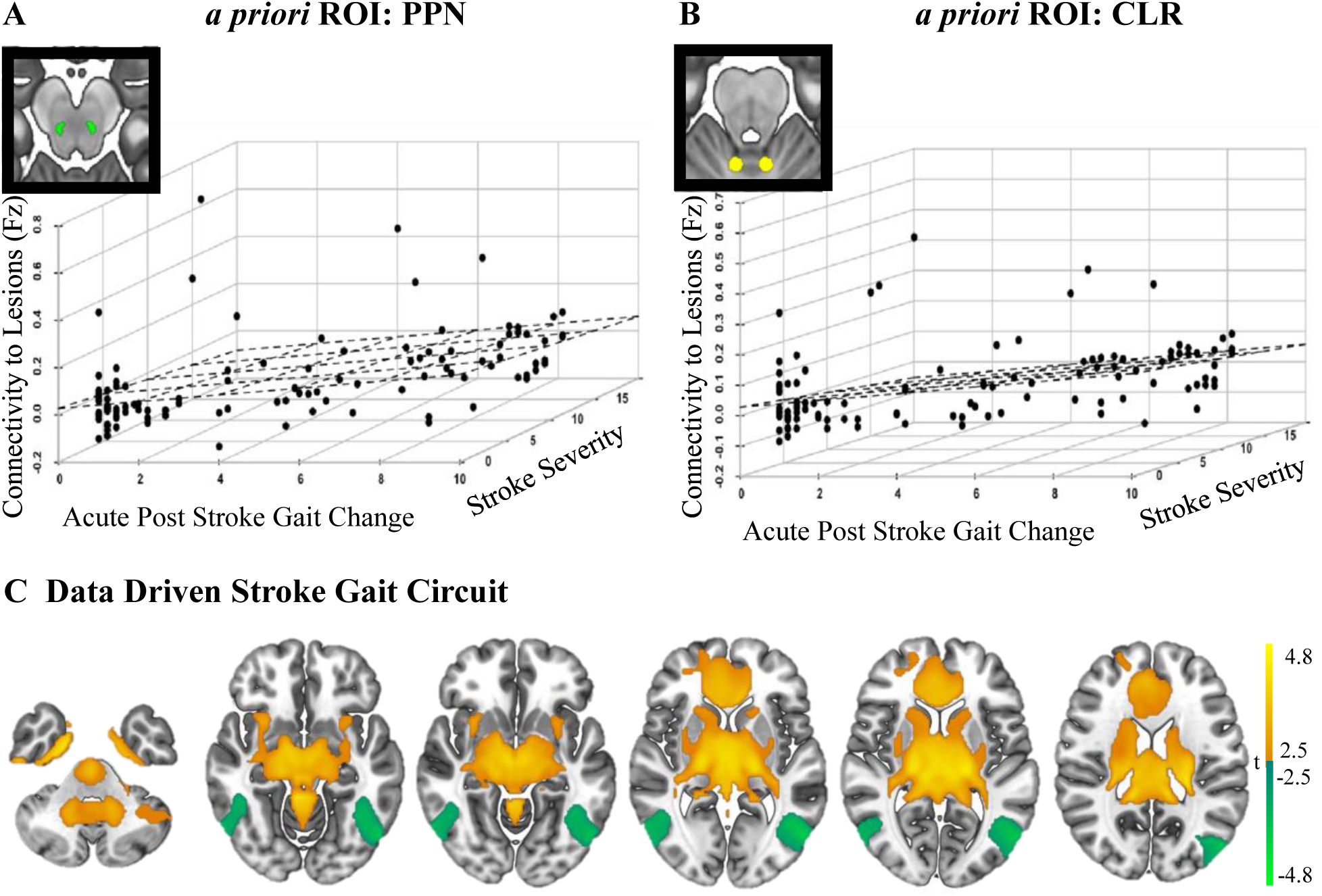
Lesions associated with post-stroke gait impairment are connected to our *a priori* ROIs and are also linked by a specific brain circuit. A, B. Connectivity between our *a priori* ROIs (PPN and CLR) and lesions correlates with clinical gait impairment in patients with stroke (PPN: r = 0.22, *p =* 0.025; CLR: r = 0.23, *p* = 0.018 after adjusting for NIHSS score). C. We also derived a data-driven lesion network which indicates brain regions significantly associated with gait impairment after correcting for stroke severity (NIHSS), at cluster-wise *pFWE* < 0.05.

### Lesion Network Mapping

Next, we performed data-driven connectivity analyses. Functional connectivity between each lesion location and the whole brain was computed using a normative connectome. Lesion connectivity was significantly associated with gait impairment, correcting for lesion size and stroke severity (cluster-based FWE-correction at *p_FWE_* < 0.05) (**Figure 2C**). Specifically, connectivity between lesion locations and a brain circuit including the bilateral parahippocampal gyrus, anterior cingulate gyrus, bilateral dorsal dentate nucleus, right cuneiform nucleus, was stronger in individuals with more gait impairment post stroke (**Supplementary Table 1**).

Connectivity between lesion locations and regions in the bilateral inferior lateral occipital cortex and left superior lateral occipital cortex was associated with less gait impairment post stroke. Note that although connectivity to our *a priori* ROIs in the PPN and CLR were significantly associated with gait impairment, neither location emerged as a peak in our data driven stroke gait circuit. The topography of our stroke gait circuit was similar after controlling for seven other post-stroke deficits including motoricity, attention, right and left motor function, spatial memory, verbal memory, and language (**Supplementary** Figure 3). Data-driven circuits for these seven post-stroke deficits failed to match the topography of our stroke gait circuit, with attention being the most similar (spatial r = −0.50) (**Supplementary** Figure 4).

### Control Analysis of Lesion Location

In contrast to the above significant results using connectivity, we did not find any lesioned voxels significantly associated with post stroke gait impairment using traditional lesion mapping approaches (again controlling for lesion size and stroke severity).

### DBS Gait Changes

After subthalamic DBS, 75 patients (60%) showed some degree of gait improvement, eight participants (6.4%) had gait impairment, while 42 subjects (33.6%) showed no change in gait. Changes in UPDRS score accounted for 58% of the variance in gait change after DBS (**Supplementary** Figure 2).

### Connectivity to A Priori Locomotor Generators

Connectivity between our 125 STN DBS sites and our two *a priori* ROIs was significantly associated with post DBS gait changes (PPN: r = 0.52, *p* = 7.84 × 10^-10^; CLR: r = 0.50, *p* = 2.19 × 10^-9^), which remained significant after correcting for both dataset and the change in total motor score (PPN: r = 0.36, *p* = 4.20 × 10^-5^; CLR: r = 0.33, *p* = 1.89 × 10^-4^) (**Figure 3A-B**).

**Figure 3.**
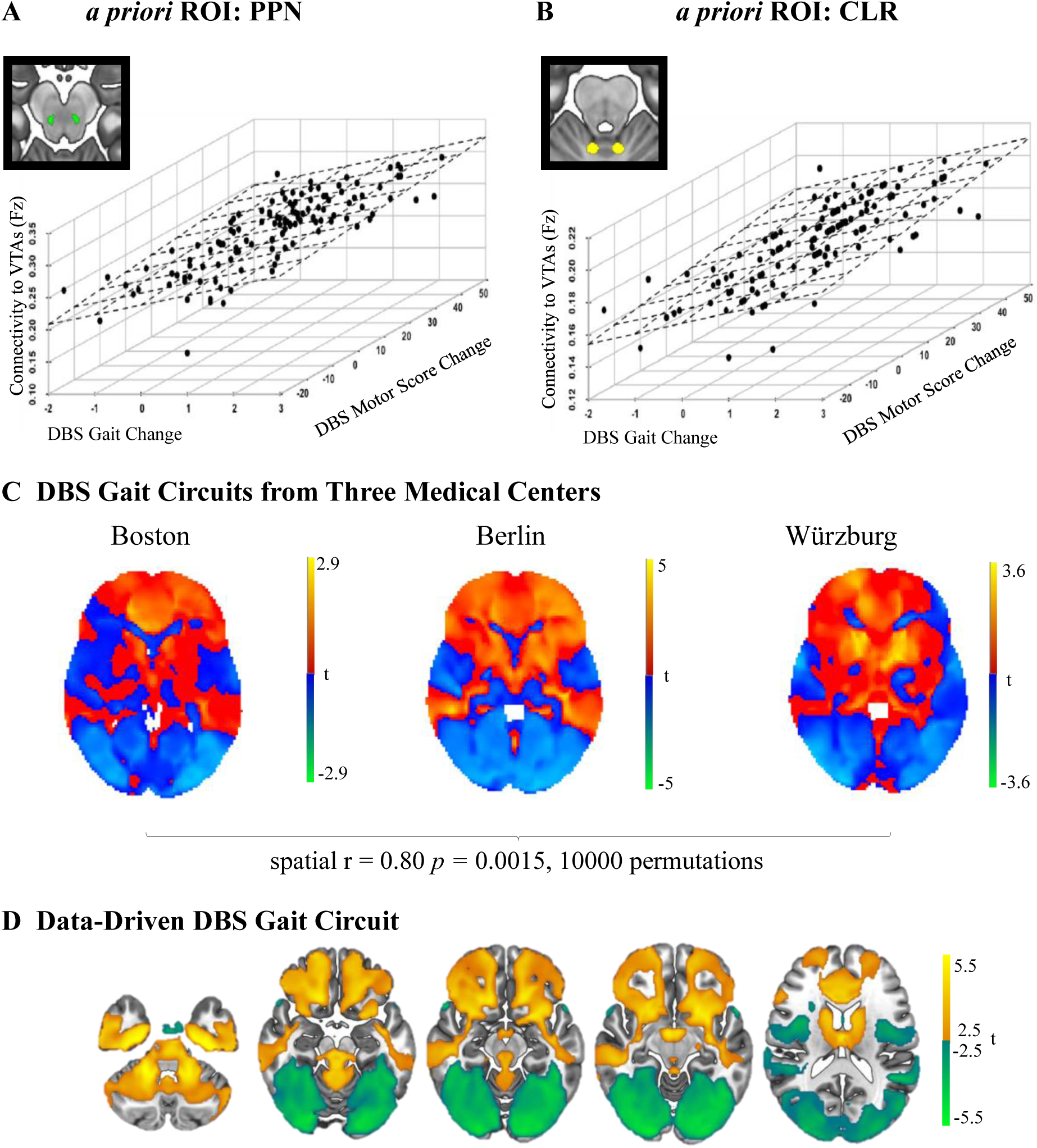
DBS sites associated with gait changes are connected to our *a priori* ROIs and are also linked by a specific brain circuit. A, B. Connectivity between our *a priori* ROIs (PPN and CLR) and DBS sites correlates with post DBS gait change in patients with PD (PPN: r = 0.36, *p* = 4.2×10^-5^; CLR: r = 0.33, *p =* 1.89×10^-4^ after adjusting for UPDRS-III and dataset). C. We derived a data-driven DBS gait circuit for each of the three DBS centers. There is high spatial correlation amongst the three circuits. D. We derived a data-driven DBS gait circuit which indicates brain regions significantly associated with gait change after controlling for change in overall motor score (UPDRS-III) and dataset, cluster-wise *pFWE* < 0.05.

### DBS Network Mapping

We derived three DBS gait circuits from each of the DBS centers (Boston n = 31; Berlin n = 51; Würzburg n = 43; **Figure 3C**). These three gait circuits demonstrated strong spatial correlation with each other (mean spatial r = 0.80, *p* = 0.0015, 10000 permutations). We also generated a data driven DBS gait circuit using the data from all three centers, and found multiple connections significantly associated with post DBS gait changes (cluster-based FWE-correction at *p_FWE_* < 0.05) (**Figure 3D**). Specifically, connectivity between DBS sites and a brain circuit including the bilateral parahippocampal gyrus, left putamen, area adjacent to left PPN, and the cerebellar vermis was associated with more gait changes after DBS (**Supplementary Table 2**).

Connectivity between DBS sites and the right cerebellum, bilateral superior parietal, and inferior occipital gyrus, and left precuneus was associated with less gait changes post DBS. Although highly significant when run as *a priori* ROIs (see above), neither the PPN nor CLR emerged as peaks within in our data-driven DBS gait circuit. The topography within our DBS gait circuit did not change after adjusting our gait circuit for other cardinal motor features in PD such as lower extremity bradykinesia, lower extremity rigidity, and postural instability (**Supplementary** Figure 5). Additionally, out of these three features, lower extremity bradykinesia contributed the most to our DBS gait circuit as it shares a high spatial topography with our DBS gait circuit (**Supplementary** Figure 6).

### Controlling for DBS Sites

In contrast to the above significant results using connectivity, traditional analyses of the stimulation sites alone failed to identify any voxels significantly associated with post DBS gait changes (again controlling for VTA size and changes in total motor score) across four different statistical approaches.

### Convergence between lesion and DBS gait circuits

Connectivity between 125 STN DBS sites and our lesion-based gait circuit was associated with DBS-induced gait changes (r = 0.43, *p* = 4.72 × 10^-7^), which remained significant after correcting for dataset and change in total motor score (r = 0.31, *p* = 4.70 × 10^-4^) (**Figure 4A**).

**Figure 4.**
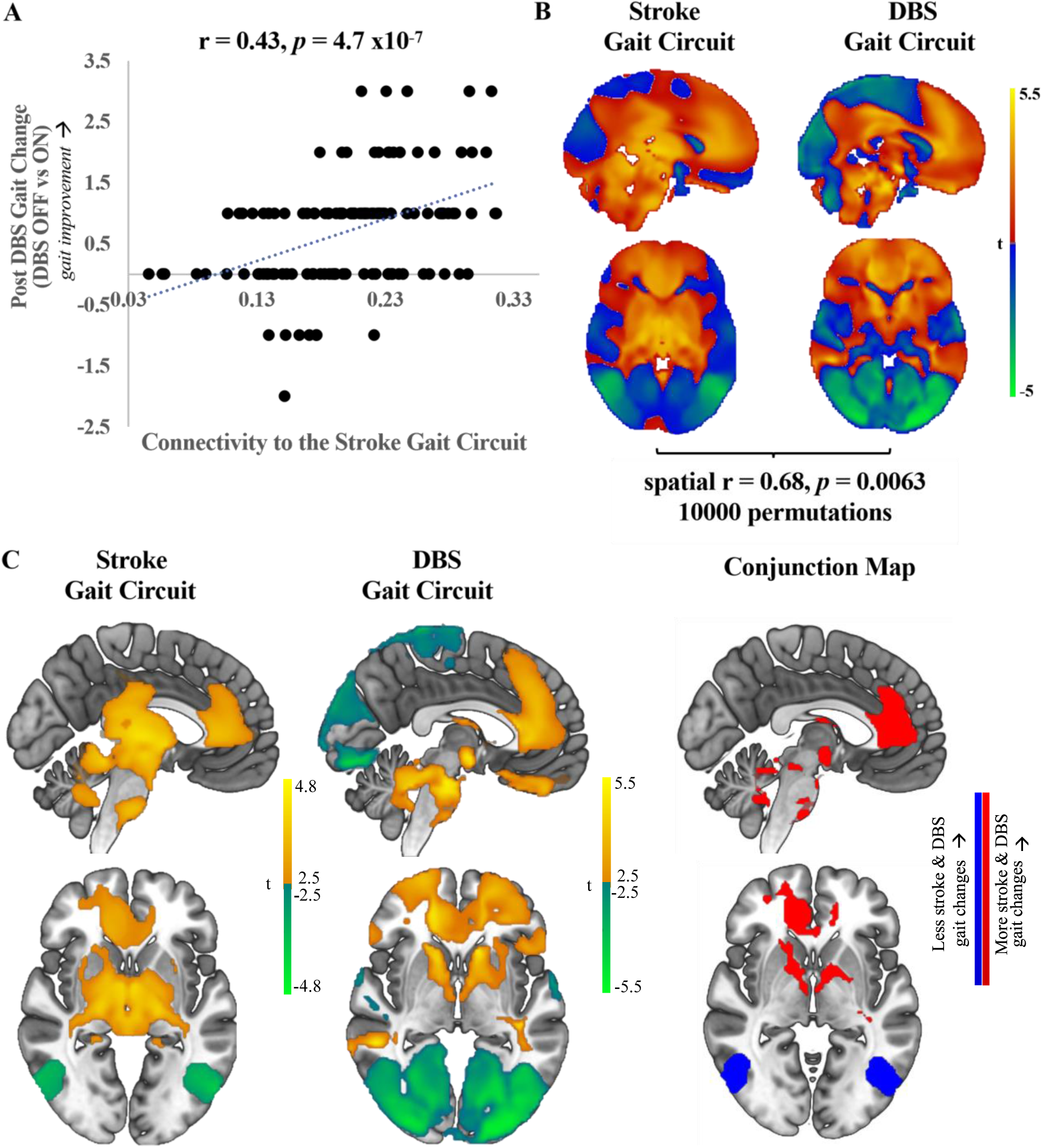
Linking stroke and DBS gait circuits. A. Connectivity between our stroke gait circuit and 125 STN DBS sites correlates with gait change after DBS (r = 0.43, *p* = 4.7 x10^-7^) which remained significant after correcting for dataset and total UPDRS-III change (r = 0.31, *p* = 4.7 x10^-4^). B. Unthresholded t maps of our stroke gait circuit and our DBS gait circuit. These two circuits show similar spatial topography and are more similar than expected by chance (spatial r = 0.68, *p* = 0.0063, 10000 permutations). C. The significant regions (cluster-wise *pFWE* <0.05) between our stroke and DBS gait circuits were overlapped and binarized to generate a conjunction map for gait.

Our two unthresholded gait circuits derived from stroke lesions and DBS sites showed similar topography (mean spatial r = 0.68, *p* = 0.0063, 10000 permutations) (**Figure 4B**).

A conjunction analysis identified multiple brain regions that were statistically significant in both the lesion and DBS circuit mapping analyses, including the left parahippocampal gyrus, right STN, bilateral pallidum, right inferior temporal gyrus, anterior and posterior cingulate cortex, cerebellar vermis, and the dentate nucleus (**Figure 4 C**). Connectivity to these regions was associated with more stroke-induced gait impairment and more DBS-induced gait changes (**Supplementary Table 3**).

## Discussion

Three major findings can be drawn from our study. First, stroke lesions associated with gait impairment can occur in multiple different brain locations, but share connectivity to a specific brain circuit, including hubs previously implicated in locomotion including the PPN and the CLR. Second, DBS sites associated with gait changes share connectivity to a specific brain circuit, including these same PPN and CLR hubs. Third, these two circuits share common topography, suggesting common neuroanatomy for gait across both lesions and DBS.

### A brain circuit for post stroke gait impairment

Prior studies of lesion location and post stroke gait impairment have implicated a variety of cortical and subcortical regions including the putamen, insula, pallidum, caudate nucleus, superior temporal gyrus, thalamus, internal and external capsule, corona radiata, corticospinal tract, superior longitudinal fasciculus, and PPN.^6^ Our results help to unify the prior literature by showing that while many different lesion locations can be associated with post-stroke gait impairment, these lesion locations share a common pattern of brain connectivity.

We found that connectivity between lesion locations and both the PPN and the CLR was associated with post stroke gait impairment. The PPN is a key component of the mesencephalic locomotor region (MLR) and is thought to initiate and maintain gait via its reciprocal connectivity to the cortex via the thalamus, basal ganglia, and spinal cord.^22,66^ The CLR is located in the middle of the cerebellar white matter (hooked bundle of Russell or uncinate fasciculus of the cerebellum) and is thought to act as a rhythm generator via fibers which decussate from the fastigial nuclei to the medullary reticular formation.^22^ Our results are consistent with a prior diffusion tensor imaging study demonstrating greater PPN structural integrity in stroke patients with better gait outcomes^67^ and another fMRI study revealing stronger activation in the CLR during imagined walking in stroke patients compared to healthy controls.^7^

Beyond these two *a priori* regions, we also derived a data-driven circuit for post-stroke gait impairment, including many of the aforementioned regions previously implicated in gait dysfunction. In particular, the parahippocampal gyrus, one of the peaks in our stroke gait circuit, is within the default mode network and is an important structure in mental navigation and topographical memory.^68–72^ It is activated during walking and has been previously linked to changes in gait speed in multiple sclerosis and freezing of gait in Parkinson’s disease.^73,74^ Furthermore, this region is activated during locomotion in a fludeoxyglucose (^18^F) (FDG)-positron emission tomography study investigating walking in older adults.^75^

There are many variables that can contribute to post stroke gait impairment,^5–7^ and many of the brain regions implicated in our stroke gait circuit have functional roles that go beyond gait.^76–78^ However, the topography of our stroke gait circuit was similar after controlling for individual variables known to contribute to gait dysfunction (such as LE weakness or attention), and data-driven maps of these variables did not match the topography of our gait circuit. These results suggest at least some specificity of this circuit to post stroke gait dysfunction.

### A brain circuit for post DBS gait change

While subthalamic DBS is an effective therapy for cardinal motor features of PD, its effects on parkinsonian gait impairment are inconsistent.^11–17^ While many clinical factors have been identified that may impact gait outcomes after STN DBS,^79^ few studies have been carried out to relate the location of stimulation to DBS gait changes.^80,81^ A small study found that stimulation in the dorsal half of the STN improved step velocity and step length of the contralateral leg compared to ventral STN stimulation.^80^ These slightly different stimulation locations within the STN could be perturbing different brain circuits.^55,82,83^

Our results also show that connectivity from the STN DBS sites to both the PPN and CLR is associated with gait changes post DBS. These results are consistent with numerous animal studies implicating the PPN in locomotion.^84–86^ Although direct targeting of the PPN with DBS has led to variable gait outcomes in PD patients,^87,88^ it is possible that indirect targeting of the PPN, via connectivity to DBS electrodes implanted in the STN, may be an important factor in gait changes after DBS. Additionally, a resting-state functional MRI study found great functional connectivity between the CLR and the supplementary motor area in PD patients with freezing of gait when compared to non-freezers.^89^

Beyond the above *a priori* regions, we also identified a data-driven DBS gait circuit. To our knowledge, this is the first study using DBS network mapping, to identify the underlying brain circuit for post DBS gait changes. This circuit has a distinctly different topography compared to our previously published post DBS motor improvement circuits^55^ and helps to unify regions previously implicated in neuroimaging correlates of parkinsonian gait including the precuneus, superior parietal lobule, inferior occipital gyrus, putamen, and cerebellar vermis.^11,90^

More importantly, gait changes after STN DBS are associated with connectivity between the stimulation site and a specific circuit of other brain regions and not one particular stimulation region. Significant peak clusters in our DBS gait circuit did not change after even adjusting for other cardinal motor features of PD including lower extremity bradykinesia, lower extremity rigidity, and postural instability. Not surprisingly, lower extremity bradykinesia circuit shares high spatial topography to our DBS gait circuit, which could indicate that gait speed is driving this network. Connectivity to our DBS gait circuit could potentially be used to guide DBS reprogramming to improve PD gait, a testable hypothesis for future work.

### A convergent brain circuit for modulating gait

One of the strongest ways to localize a symptom in the human brain is to use two complementary but independent causal sources of data, such as lesions and DBS locations.^32^ Here, we applied this approach for gait and identified clear convergence despite many differences in gait dysfunction in stroke and PD. Our convergent brain circuit was uncovered despite heterogeneity in stroke locations, stimulation sites, gait phenomenology (hemiparetic versus parkinsonian gait), and clinical gait metrics (combined walking index versus UDPRS gait subscore). This convergence across different sources of data (and presumably different etiologies for gait change) increases confidence that our circuit is indeed related to gait, rather than a co-morbid variable associated with stroke or PD alone. Note that this convergence may be unexpected, as most would assume different mechanism and thus different brain circuits for stroke and PD gait impairment. However, this convergent is consistent with multiple recent studies in other neuropsychiatric disorders, which showed convergence between lesions and DBS.^25–31^

Our convergent brain circuit for gait using lesion and DBS aligns with prior work on neuroimaging correlates of gait. Brain regions implicated in both our convergent circuit and prior neuroimaging studies include the anterior and posterior cingulate cortex, inferior temporal gyrus, STN, globus pallidum, bilateral thalamus, cerebellar vermis, and pons.^22,76,91,92^ Interestingly, the topography of these regions have been implicated in dopaminergic and cholinergic neural pathways which have been previously shown to play a major role in parkinsonian gait impairment.^93,94^

### Limitations

This study has four important limitations. First, our analyses were all retrospective and clinical relevance of these gait circuits has yet to be tested prospectively. Second, while we accounted for stroke severity and total motor changes after subthalamic DBS in our derivation of our gait circuits, we did not factor freezing of gait, polyneuropathy, cerebellar ataxia, sensory deficits, or musculoskeletal abnormalities in our analyses which could all impact gait phenomenology.

Third, we used two different clinical rating scales to define gait impairment (combined walking index versus UPDRS gait scores) and it is unclear whether our results hold after using objective gait metrics such as gait speed or stride-to-stride variability. Fourth, all analyses were performed using a normative functional connectome instead of a disease-specific connectome or patient-specific connectivity. However, prior research indicate highly similar functional connectivity results when using the normative versus PD connectome^55^ and when using a normative connectome versus patient-specific connectivity.^95^ In the end, despite the aforementioned sources of heterogeneity and noise, we were still able to find a convergent gait circuit across modalities.

### Conclusions

In conclusion, by combining stroke lesions and DBS sites, we found a common brain circuit associated with gait impairment following stroke and gait changes after subthalamic DBS. Our convergent gait circuit may hold promise as a therapeutic target to improve gait impairment across disorders.

## Data Availability

All data produced in the present study are available upon reasonable request to the authors.

## Acknowledgements

The authors would like to thank the patients who participated in this study and the researchers for publicly sharing their results; and the lab members of the NimLab at Beth Israel Deaconess Medical Center and the Center for Brain Circuit Therapeutics at Brigham and Women’s Hospital who helped develop some of the code employed in this study. L.L. would like to thank the Parkinson Study Group for awarding her with the Mentored Clinical Research Award. F.S. is supported by the NIH and the American Epilepsy Society. A.H. is supported by the German Research Foundation (Deutsche Forschungsgemeinschaft, 424778381–TRR 295), Deutsches Zentrum fur Luft-und Raumfahrt (DynaSti Grant within the EU Joint Programme Neurodegenerative Disease Research, JPND), the NIH (R01 13478451, 1R01NS127892-01, 2R01 MH113929, and UM1NS132358) as well as the New Venture Fund (FFOR Seed Grant). M. R. is supported by the Deutsche Forschungsgemeinschaft (DFG, German Research Foundation): Project-ID 424778381–TRR 295; Project B07. A.A.K. is supported by the Deutsche Forschungsgemeinschaft (DFG, German Research Foundation). R.L.A. is supposed by a grant from Medtronic. M.D.F. is supported by grants from the NIH (R01MH113929, R21MH126271, R56AG069086, R21NS123813, R01NS127892, R01MH130666, and UM1NS132358), Neuronetics, the Kaye Family Research Endowment, the Ellison/Baszucki Family Foundation, and the Manley Family.

## Author Contributions

Study concept and design: all authors; data acquisition and analysis: all authors; drafting the text and figures: L.L, M.D.F.

## References

1. Chua KSG, Chee J, Wong CJ, et al. A pilot clinical trial on a Variable Automated Speed and Sensing Treadmill (VASST) for hemiparetic gait rehabilitation in stroke patients [Internet]. Front Neurosci 2015;9 [cited 2020 Oct 31] Available from: https://www.ncbi.nlm.nih.gov/pmc/articles/PMC4498099/

2. Hill K, Ellis P, Bernhardt J, et al. Balance and mobility outcomes for stroke patients: a comprehensive audit. Aust J Physiother 1997;43(3):173–180.

3. Gresham GE, Fitzpatrick TE, Wolf PA, et al. Residual disability in survivors of stroke--the Framingham study. N Engl J Med 1975;293(19):954–956.

4. Portelli R, Lowe D, Irwin P, et al. Institutionalization after stroke. Clinical Rehabilitation; London 2005;19(1):97–108.

5. Allali G, Blumen HM, Devanne H, et al. Brain imaging of locomotion in neurological conditions. Neurophysiologie Clinique 2018;48(6):337–359.

6. Perry MK, Peters DM. Neural correlates of walking post-stroke: neuroimaging insights from the past decade. Exp Brain Res 2021;239(12):3439–3446.

7. Boyne P, Doren S, Scholl V, et al. Functional magnetic resonance brain imaging of imagined walking to study locomotor function after stroke. Clinical Neurophysiology 2021;132(1):167–177.

8. Giladi N, Treves TA, Simon ES, et al. Freezing of gait in patients with advanced Parkinson’s disease. J Neural Transm (Vienna) 2001;108(1):53–61.

9. Fasano A, Canning CG, Hausdorff JM, et al. Falls in Parkinson’s disease: A complex and evolving picture. Mov Disord 2017;32(11):1524–1536.

10. Muslimovic D, Post B, Speelman JD, et al. Determinants of disability and quality of life in mild to moderate Parkinson disease. Neurology 2008;70(23):2241–2247.

11. Mirelman A, Bonato P, Camicioli R, et al. Gait impairments in Parkinson’s disease. The Lancet Neurology 2019;18(7):697–708.

12. Mei S, Eisinger RS, Hu W, et al. Three-Year Gait and Axial Outcomes of Bilateral STN and GPi Parkinson’s Disease Deep Brain Stimulation. Front Hum Neurosci 2020;14:1.

13. Liu W, McIntire K, Kim SH, et al. Quantitative assessments of the effect of bilateral subthalamic stimulation on multiple aspects of sensorimotor function for patients with Parkinson’s disease. Parkinsonism & Related Disorders 2005;11(8):503–508.

14. Liu W, McIntire K, Kim SH, et al. Bilateral subthalamic stimulation improves gait initiation in patients with Parkinson’s disease. Gait & Posture 2006;23(4):492–498.

15. Vallabhajosula S, Haq IU, Hwynn N, et al. Low-frequency Versus High-frequency Subthalamic Nucleus Deep Brain Stimulation on Postural Control and Gait in Parkinson’s Disease: A Quantitative Study. Brain Stimulation 2015;8(1):64–75.

16. Chastan N, Do MC, Bonneville F, et al. Gait and balance disorders in Parkinson’s disease: Impaired active braking of the fall of centre of gravity. Movement Disorders 2009;24(2):188–195.

17. Crenna P, Carpinella I, Rabuffetti M, et al. Impact of subthalamic nucleus stimulation on the initiation of gait in Parkinson’s disease. Exp Brain Res 2006;172(4):519–532.

18. Fleury V, Pollak P, Gere J, et al. Subthalamic stimulation may inhibit the beneficial effects of levodopa on akinesia and gait. Mov Disord 2016;31(9):1389–1397.

19. Pötter-Nerger M, Volkmann J. Deep brain stimulation for gait and postural symptoms in Parkinson’s disease. Mov Disord 2013;28(11):1609–1615.

20. Thevathasan W, Debu B, Aziz T, et al. Pedunculopontine nucleus deep brain stimulation in Parkinson’s disease: A clinical review. Movement Disorders 2018;33(1):10–20.

21. Hausdorff JM, Alexander NB. Gait disordersL: evaluation and management. Boca Raton: Taylor & Francis; 2005.

22. Takakusaki K. Neurophysiology of gait: From the spinal cord to the frontal lobe. Movement Disorders 2013;28(11):1483–1491.

23. Jahn K, Deutschländer A, Stephan T, et al. Supraspinal locomotor control in quadrupeds and humans. Prog Brain Res 2008;171:353–362.

24. Weiss PH, Herzog J, Pötter-Nerger M, et al. Subthalamic nucleus stimulation improves Parkinsonian gait via brainstem locomotor centers. Movement Disorders 2015;30(8):1121–1125.

25. Joutsa J, Horn A, Hsu J, Fox MD. Localizing parkinsonism based on focal brain lesions. Brain 2018;141(8):2445–2456.

26. Corp DT, Joutsa J, Darby RR, et al. Network localization of cervical dystonia based on causal brain lesions. Brain 2019;142(6):1660–1674.

27. Okromelidze L, Tsuboi T, Eisinger RS, et al. Functional and Structural Connectivity Patterns Associated with Clinical Outcomes in Deep Brain Stimulation of the Globus Pallidus Internus for Generalized Dystonia. AJNR Am J Neuroradiol 2020;41(3):508– 514.

28. Siddiqi SH, Schaper F, Hsu J, et al. Convergent causal mapping of human neuropsychiatric symptoms using brain stimulation and brain lesions. Nature Human Behaviour [date unknown];

29. Reich MM, Hsu J, Ferguson M, et al. A brain network for deep brain stimulation induced cognitive decline in Parkinson’s disease. Brain 2022;awac012.

30. Ganos C, Al-Fatly B, Fischer J-F, et al. A neural network for tics: insights from causal brain lesions and deep brain stimulation. Brain 2022;awac009.

31. Schaper FLWVJ, Nordberg J, Cohen AL, et al. Mapping Lesion-Related Epilepsy to a Human Brain Network. JAMA Neurol 2023;

32. Siddiqi SH, Kording KP, Parvizi J, Fox MD. Causal mapping of human brain function. Nat Rev Neurosci 2022;23(6):361–375.

33. Boes AD, Prasad S, Liu H, et al. Network localization of neurological symptoms from focal brain lesions. Brain 2015;138(10):3061–3075.

34. Darby RR, Laganiere S, Pascual-Leone A, et al. Finding the imposter: brain connectivity of lesions causing delusional misidentifications. Brain 2017;140(2):497–507.

35. Darby RR, Horn A, Cushman F, Fox MD. Lesion network localization of criminal behavior. Proc Natl Acad Sci U S A 2018;115(3):601–606.

36. Fasano A, Laganiere SE, Lam S, Fox MD. Lesions causing freezing of gait localize to a cerebellar functional network: Lesion Network Mapping and FOG. Ann Neurol. 2017;81(1):129–141.

37. Fischer DB, Boes AD, Demertzi A, et al. A human brain network derived from coma-causing brainstem lesions. Neurology 2016;87(23):2427–2434.

38. Joutsa J, Shih LC, Fox MD. Mapping holmes tremor circuit using the human brain connectome. Ann Neurol 2019;86(6):812–820.

39. Laganiere S, Boes AD, Fox MD. Network localization of hemichorea-hemiballismus. Neurology 2016;86(23):2187–2195.

40. Padmanabhan JL, Cooke D, Joutsa J, et al. A Human Depression Circuit Derived From Focal Brain Lesions. Biological Psychiatry 2019;86(10):749–758.

41. Fox MD. Mapping Symptoms to Brain Networks with the Human Connectome. N Engl J Med 2018;379(23):2237–2245.

42. Joutsa J, Shih LC, Horn A, et al. Identifying therapeutic targets from spontaneous beneficial brain lesions. Annals of Neurology 2018;84(1):153–157.

43. Corbetta M, Ramsey L, Callejas A, et al. Common Behavioral Clusters and Subcortical Anatomy in Stroke. Neuron 2015;85(5):927–941.

44. Ortiz GA, Sacco RL. National Institutes of Health Stroke Scale (NIHSS) [Internet]. In: Wiley StatsRef: Statistics Reference Online. American Cancer Society; 2014[cited 2020 Nov 1] Available from: http://onlinelibrary.wiley.com/doi/abs/10.1002/9781118445112.stat06823

45. Perry J, Garrett M, Gronley JK, Mulroy SJ. Classification of walking handicap in the stroke population. Stroke 1995;26(6):982–989.

46. Keith RA, Granger CV, Hamilton BB, Sherwin FS. The functional independence measure: a new tool for rehabilitation. Adv Clin Rehabil 1987;1:6–18.

47. Alho ATDL, Hamani C, Alho EJL, et al. Magnetic resonance diffusion tensor imaging for the pedunculopontine nucleus: proof of concept and histological correlation. Brain Struct Funct 2017;222(6):2547–2558.

48. Boyne P, Maloney T, DiFrancesco M, et al. Resting-state functional connectivity of subcortical locomotor centers explains variance in walking capacity. Hum Brain Mapp 2018;39(12):4831–4843.

49. Yeo BTT, Krienen FM, Sepulcre J, et al. The organization of the human cerebral cortex estimated by intrinsic functional connectivity. J Neurophysiol 2011;106(3):1125– 1165.

50. Boes AD, Prasad S, Liu H, et al. Network localization of neurological symptoms from focal brain lesions. Brain 2015;138(Pt 10):3061–3075.

51. Joutsa J, Shih LC, Horn A, et al. Identifying therapeutic targets from spontaneous beneficial brain lesions. Ann Neurol 2018;84(1):153–157.

52. Winkler AM, Ridgway GR, Webster MA, et al. Permutation inference for the general linear model. NeuroImage 2014;92:381–397.

53. The MathWorks Inc.: MATLAB and Statistics Toolbox Release 2015b. 2015;

54. Rorden C, Karnath H-O, Bonilha L. Improving lesion-symptom mapping. J Cogn Neurosci 2007;19(7):1081–1088.

55. Horn A, Reich M, Vorwerk J, et al. Connectivity Predicts deep brain stimulation outcome in Parkinson disease: DBS Outcome in PD. Ann Neurol. 2017;82(1):67–78.

56. Fahn S, Elton, RL. Unified Parkinson’s Disease Rating Scale. In: Recent Developments in Parkinson’s Disease. Florham Park, NJ: Macmillan Healthcare Information; 1987 p. 153–163, 293–304.

57. Goetz CG, Tilley BC, Shaftman SR, et al. Movement Disorder Society-sponsored revision of the Unified Parkinson’s Disease Rating Scale (MDS-UPDRS): scale presentation and clinimetric testing results. Mov Disord 2008;23(15):2129–2170.

58. Horn A, Kühn AA. Lead-DBS: a toolbox for deep brain stimulation electrode localizations and visualizations. Neuroimage 2015;107:127–135.

59. Horn A, Li N, Dembek TA, et al. Lead-DBS v2: Towards a comprehensive pipeline for deep brain stimulation imaging. NeuroImage 2019;184:293–316.

60. Avants BB, Epstein CL, Grossman M, Gee JC. Symmetric diffeomorphic image registration with cross-correlation: evaluating automated labeling of elderly and neurodegenerative brain. Med Image Anal 2008;12(1):26–41.

61. Siddiqi SH, Schaper FLWVJ, Horn A, et al. Brain stimulation and brain lesions converge on common causal circuits in neuropsychiatric disease. Nat Hum Behav 2021;

62. Butson CR, Cooper SE, Henderson JM, et al. Probabilistic analysis of activation volumes generated during deep brain stimulation. Neuroimage 2011;54(3):2096–2104.

63. Cheung T, Noecker AM, Alterman RL, et al. Defining a therapeutic target for pallidal deep brain stimulation for dystonia. Ann Neurol 2014;76(1):22–30.

64. Reich MM, Horn A, Lange F, et al. Probabilistic mapping of the antidystonic effect of pallidal neurostimulation: a multicentre imaging study. Brain 2019;142(5):1386– 1398.

65. Dembek TA, Roediger J, Horn A, et al. Probabilistic sweet spots predict motor outcome for deep brain stimulation in Parkinson disease. Annals of Neurology 2019;86(4):527– 538.

66. Benarroch EE. Pedunculopontine nucleus: Functional organization and clinical implications. Neurology 2013;80(12):1148–1155.

67. Yeo SS, Ahn SH, Choi BY, et al. Contribution of the Pedunculopontine Nucleus on Walking in Stroke Patients. ENE 2011;65(6):332–337.

68. Berthoz A. Parietal and hippocampal contribution to topokinetic and topographic memory. Philos Trans R Soc Lond B Biol Sci 1997;352>(1360):1437–1448.

69. Maguire EA, Frith CD, Burgess N, et al. Knowing where things are parahippocampal involvement in encoding object locations in virtual large-scale space. J Cogn Neurosci 1998;10(1):61–76.

70. Maguire EA, Burke T, Phillips J, Staunton H. Topographical disorientation following unilateral temporal lobe lesions in humans. Neuropsychologia 1996;34(10):993– 1001.

71. Mellet E, Briscogne S, Tzourio-Mazoyer N, et al. Neural correlates of topographic mental exploration: the impact of route versus survey perspective learning. Neuroimage 2000;12(5):588–600.

72. Habib M, Sirigu A. Pure topographical disorientation: a definition and anatomical basis. Cortex 1987;23(1):73–85.

73. Bollaert RE, Poe K, Hubbard EA, et al. Associations of functional connectivity and walking performance in multiple sclerosis. Neuropsychologia 2018;117:8–12.

74. Jin C, Qi S, Teng Y, et al. Altered Degree Centrality of Brain Networks in Parkinson’s Disease With Freezing of Gait: A Resting-State Functional MRI Study. Front Neurol 2021;12:743135.

75. la Fougère C, Zwergal A, Rominger A, et al. Real versus imagined locomotion: a [18F]-FDG PET-fMRI comparison. Neuroimage 2010;50(4):1589–1598.

76. Maiti B, Rawson KS, Tanenbaum AB, et al. Functional Connectivity of Vermis Correlates with Future Gait Impairments in Parkinson’s Disease [Internet]. Movement Disorders [date unknown];n/a(n/a)[cited 2021 Jul 17] Available from: http://movementdisorders.onlinelibrary.wiley.com/doi/abs/10.1002/mds.28684

77. Rolls ET. The cingulate cortex and limbic systems for emotion, action, and memory. Brain Struct Funct 2019;224(9):3001–3018.

78. Peng X, Burwell RD. Beyond the hippocampus: The role of parahippocampal-prefrontal communication in context-modulated behavior. Neurobiol Learn Mem 2021;185:107520.

79. Russmann H, Ghika J, Villemure J-G, et al. Subthalamic nucleus deep brain stimulation in Parkinson disease patients over age 70 years. Neurology 2004;63(10):1952–1954.

80. Johnsen EL, Sunde N, Mogensen PH, Ostergaard K. MRI verified STN stimulation site--gait improvement and clinical outcome. Eur J Neurol 2010;17(5):746–753.

81. Schott FP, Gulberti A, Pinnschmidt HO, et al. Subthalamic Deep Brain Stimulation Lead Asymmetry Impacts the Parkinsonian Gait Disorder. Front Hum Neurosci 2022;16:788200.

82. Doya K. Complementary roles of basal ganglia and cerebellum in learning and motor control. Curr Opin Neurobiol 2000;10(6):732–739.

83. Horn A, Fox MD. Opportunities of connectomic neuromodulation. Neuroimage 2020;221:117180.

84. Brudzynski SM, Houghton PE, Brownlee RD, Mogenson GJ. Involvement of neuronal cell bodies of the mesencephalic locomotor region in the initiation of locomotor activity of freely behaving rats. Brain Res Bull 1986;16(3):377–381.

85. Milner KL, Mogenson GJ. Electrical and chemical activation of the mesencephalic and subthalamic locomotor regions in freely moving rats. Brain Res 1988;452(1–2):273– 285.

86. Jenkinson N, Nandi D, Miall RC, et al. Pedunculopontine nucleus stimulation improves akinesia in a Parkinsonian monkey. Neuroreport 2004;15(17):2621–2624.

87. Nowacki A, Galati S, Ai-Schlaeppi J, et al. Pedunculopontine nucleus: An integrative view with implications on Deep Brain Stimulation. Neurobiol Dis 2019;128:75–85.

88. Chambers NE, Lanza K, Bishop C. Pedunculopontine Nucleus Degeneration Contributes to Both Motor and Non-Motor Symptoms of Parkinson’s Disease. Front Pharmacol 2019;10:1494.

89. Fling BW, Cohen RG, Mancini M, et al. Functional reorganization of the locomotor network in Parkinson patients with freezing of gait. PLoS One 2014;9(6):e100291.

90. Peterson DS, Horak FB. Neural Control of Walking in People with Parkinsonism. Physiology 2016;31(2):95–107.

91. McGough EL, Kelly VE, Weaver KE, et al. Limbic and Basal Ganglia Neuroanatomical Correlates of Gait and Executive Function. Am J Phys Med Rehabil 2018;97(4):229– 235.

92. Byun S, Lee HJ, Kim JS, et al. Exploring shared neural substrates underlying cognition and gait variability in adults without dementia. Alzheimers Res Ther 2023;15(1):206.

93. Bohnen NI, Kanel P, Koeppe RA, et al. Regional cerebral cholinergic nerve terminal integrity and cardinal motor features in Parkinson’s disease. Brain Commun 2021;3(2):fcab109.

94. Stuart S, Morris R, Giritharan A, et al. Prefrontal Cortex Activity and Gait in Parkinson’s Disease With Cholinergic and Dopaminergic Therapy [Internet]. Movement Disorders [date unknown];n/a(n/a)[cited 2020 Nov 1] Available from: http://onlinelibrary.wiley.com/doi/abs/10.1002/mds.28214

95. Wang Q, Akram H, Muthuraman M, et al. Normative vs. patient-specific brain connectivity in deep brain stimulation. NeuroImage 2021;224:117307.

