## Supplementary Figure 1 for "A Human Gait Circuit Derived from Brain Lesions and Deep Brain Stimulation"

**Supplementary Figure 1. Different lesions produce different severity of gait impairment.** A. As expected, there is a correlation between gait impairment and overall stroke severity (r = 0.71, $p < 0.00001$). For visual clarity, post stroke gait scores are displayed as 10 minus total walk score so that higher values indicate more gait impairment. B. However, lesions can result in significant gait impairment despite low stroke severity. C. Conversely, lesions can result in no gait impairment despite high stroke severity. D, E. Lesions that appear in a similar anatomical location can be associated with different degrees of gait impairment (compare B to D and C to E). Individuals shown in B-E are denoted by letters on the scatter plot.
NIHSS = National Institutes of Health Stroke Scale.

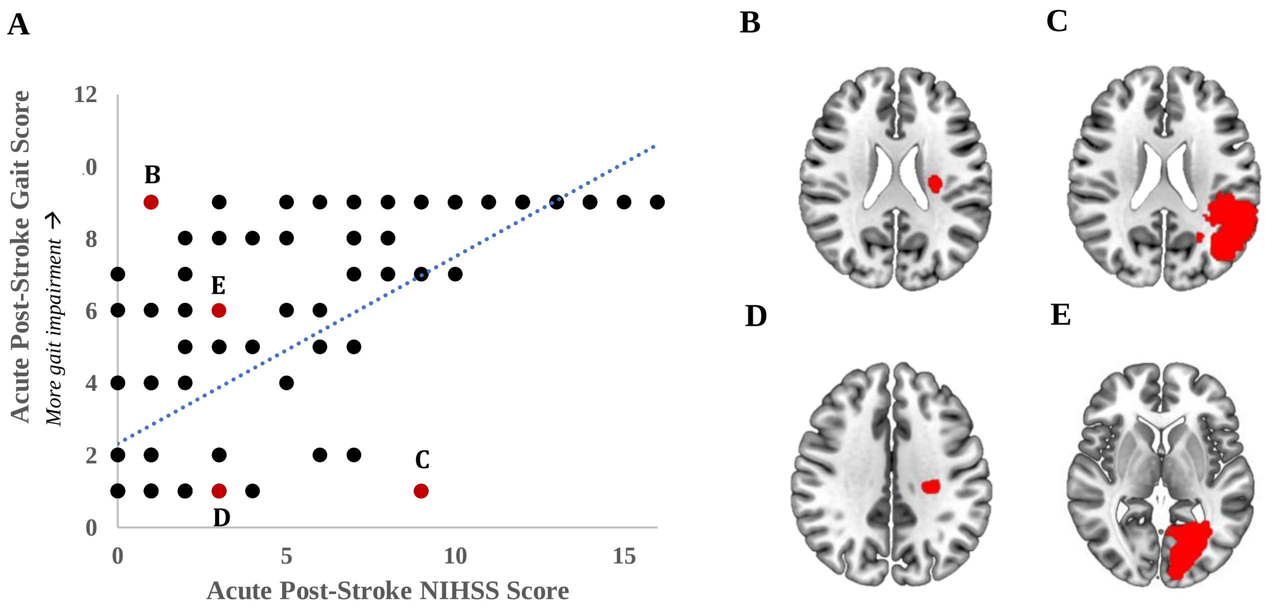
