## Supplementary Figure 2 for "A Human Gait Circuit Derived from Brain Lesions and Deep Brain Stimulation"

**Supplementary Figure 2. Different DBS sites produce different amounts of gait change.**
A. As expected, there is a correlation between change in gait score and change in total motor score after bilateral STN DBS ($r = 0.58$, $p < 1x10^{-5}$). However, VTAs in similar brain locations can result either in gait improvement, no gait change, or gait deterioration after bilateral STN DBS. B-E. DBS electrode localizations (shown in red) along with its corresponding VTAs (shown in purple) (B, C) for two subjects who had gait improvement and (D, E) two subjects who had gait deterioration after DBS to the STN (structure shown in orange).
Individuals shown in B-E are denoted by letters on the scatter plot.

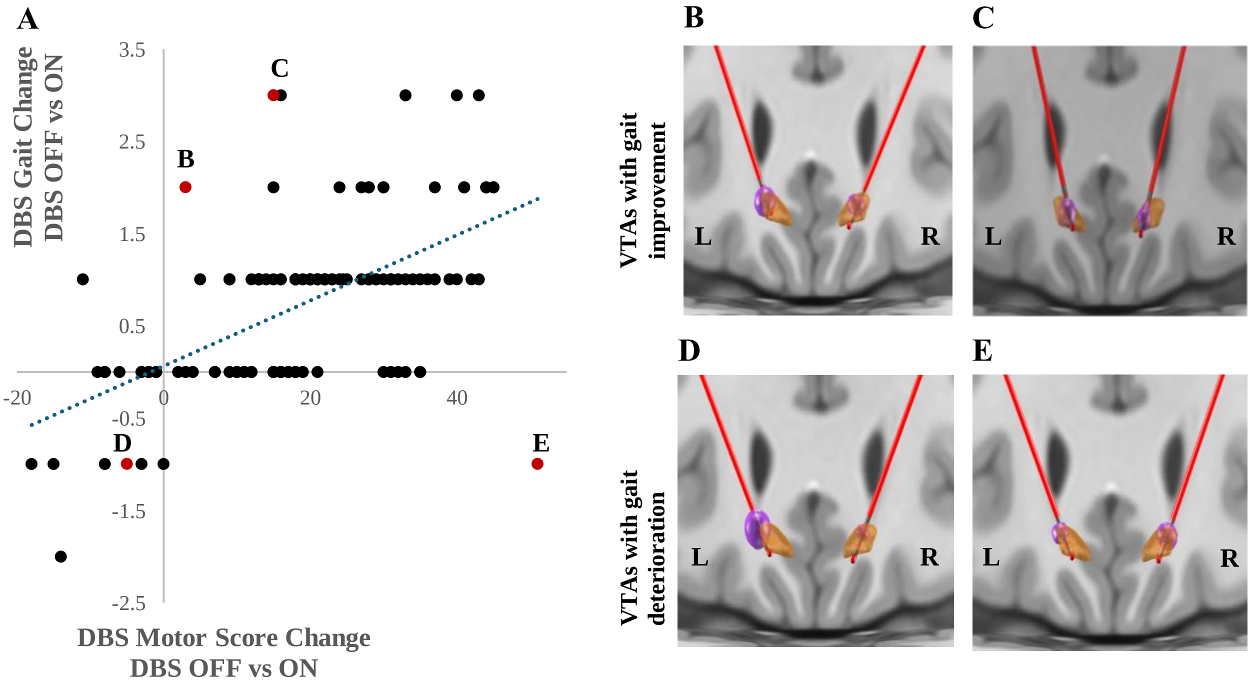
