## Supplementary Table 1 for "A Human Gait Circuit Derived from Brain Lesions and Deep Brain Stimulation"

**Supplementary Table 1.**
Coordinates of peak clusters from our stroke gait circuit.

| ***Cluster-wise $p_{FWE}$*** | ***T* value** | **Voxels** | **X (mm)** | **Y (mm)** | **Z (mm)** | **Region** |
|---|---|---|---|---|---|---|
| < 0.05 | 4.55 | 5288 | -20 | -22 | 8 | Left Ventral Posterior Lateral Nucleus of the Thalamus (L VPl) |
| | 4.52 | 5288 | 20 | -20 | 12 | Right Ventral Posterior Lateral Nucleus of the Thalamus (R VPl) |
| | 4.46 | 196 | 20 | -4 | -38 | Right Parahippocampal Gyrus |
| | 4.25 | 5288 | 6 | -16 | -8 | Right Red nucleus |
| | 4.14 | 5288 | -4 | -16 | -8 | Left Red nucleus |
| | 3.79 | 40 | -26 | -14 | -36 | Left Parahippocampal Gyrus |
| | 3.74 | 550 | 2 | 36 | 4 | Anterior Cingulate Gyrus |
| | 3.52 | 2 | -14 | -50 | -38 | Left Dorsal Dentate nucleus |
| | 3.52 | 2 | 16 | -56 | -36 | Right Dorsal Dentate nucleus |
| | 3.76 | 5288 | 4 | -28 | -12 | Right Cuneiform Nucleus |

| ***Cluster-wise $p_{FWE}$*** | ***T* value** | **Voxels** | **X (mm)** | **Y (mm)** | **Z (mm)** | **Region** |
|---|---|---|---|---|---|---|
| < 0.05 | -4.00 | 745 | -54 | -68 | 2 | Left Inferior Lateral Occipital Cortex |
| | -3.87 | 484 | -50 | -76 | 20 | Left Superior Lateral Occipital Cortex |
| | -3.62 | 103 | 58 | -64 | -4 | Right Inferior Lateral Occipital Cortex |
