## Supplementary Table 2 for "A Human Gait Circuit Derived from Brain Lesions and Deep Brain Stimulation"

**Supplementary Table 2.**
Coordinates of peak clusters from our DBS gait circuit.
DBS = deep brain stimulation.

| ***Cluster-wise $p_{FWE}$*** | **T value** | **Voxels** | **X (mm)** | **Y (mm)** | **Z (mm)** | **Region** |
|---|---|---|---|---|---|---|
| <0.05 | 5.23 | 1004 | -34 | -18 | -34 | Left Fusiform Gyrus |
| | 5.17 | 1004 | -28 | -14 | -32 | Left Parahippocampal Gyrus |
| | 5.13 | 2282 | 26 | -12 | -34 | Right Parahippocampal Gyrus |
| | 5.11 | 2282 | 44 | -16 | -38 | Right Fusiform Gyrus |
| | 4.78 | 515 | -22 | 12 | -10 | Left Putamen |
| | 4.75 | 581 | 16 | 24 | -18 | Right Superior Orbital Gyrus |
| | 4.67 | 515 | -16 | 24 | -14 | Left Superior Orbital Gyrus |
| | 4.38 | 2282 | -8 | -30 | -20 | Left PPN |
| | 4.28 | 3 | 0 | -46 | -34 | Cerebellar Vermis |

| ***Cluster-wise $p_{FWE}$*** | ***T* value** | **Voxels** | **X (mm)** | **Y (mm)** | **Z (mm)** | **Region** |
|---|---|---|---|---|---|---|
| <0.05 | -4.64 | 607 | 26 | -84 | -18 | Right Cerebellum (Crus I) |
| | -4.62 | 40 | -20 | -70 | 64 | Left Superior Parietal Lobule |
| | -4.55 | 607 | 24 | -94 | -12 | Right Lingual Gyrus |
| | -4.53 | 607 | 32 | -92 | -6 | Right Inferior Occipital Gyrus |
| | -4.43 | 1005 | -16 | -94 | -14 | Left Lingual Gyrus |
| | -4.43 | 1005 | -28 | -92 | -8 | Left Inferior Occipital Gyrus |
| | -4.38 | 49 | 24 | -58 | 72 | Right Superior Parietal Lobule |
| | -4.37 | 16 | 28 | -84 | 46 | Right Superior Occipital Gyrus |
| | -4.33 | 17 | -12 | -56 | 76 | Left Precuneus |
