## Supplementary Table 3 for "A Human Gait Circuit Derived from Brain Lesions and Deep Brain Stimulation"

**Supplementary Table 3. Coordinates of peak clusters from our conjunction analysis.**
Peak clusters significantly associated with both gait changes post stroke and post DBS.
DBS = deep brain stimulation.

| ***Cluster-wise $p_{FWE}$*** | ***T* value** | **Voxels** | **X (mm)** | **Y (mm)** | **Z (mm)** | **Region** |
|---|---|---|---|---|---|---|
| < 0.05 | 4.31 | 186 | -24 | -12 | -34 | Left Parahippocampal Gyrus |
| | 4.24 | 210 | 6 | -10 | -8 | Right Subthalamic Nucleus |
| | 4.18 | 72 | 52 | -8 | -42 | Right Inferior Temporal Gyrus |
| | 3.93 | 742 | 2 | 26 | 10 | Anterior Cingulate Cortex |
| | 3.92 | 37 | 0 | -42 | -12 | Cerebellar Vermis |
| | 3.79 | 19 | 14 | 4 | 0 | Right Pallidum |
| | 3.74 | 30 | 16 | -50 | -34 | Dentate nucleus |
| | 3.68 | 86 | -8 | -16 | 18 | Left Ventrolateral Posterior Nucleus of the Thalamus (L VLp) |
| | 3.65 | 210 | -12 | -2 | 0 | Left Pallidum |
| | 3.63 | 86 | 10 | -14 | 18 | Right Ventrolateral Posterior Nucleus of the Thalamus (R VLp) |
| | 3.54 | 30 | 0 | -26 | -40 | Ventral Pons |
| | 3.53 | 2 | -4 | -32 | 22 | Posterior Cingulate Cortex |

| ***Cluster-wise $p_{FWE}$*** | ***T* value** | **Voxels** | **X (mm)** | **Y (mm)** | **Z (mm)** | **Region** |
|---|---|---|---|---|---|---|
| < 0.05 | -4.18 | 139 | -24 | -66 | 66 | Left Superior Parietal Lobule |
| | -3.8 | 432 | 46 | -62 | -12 | Right Inferior Temporal Gyrus |
| | -3.64 | 432 | -46 | -48 | -16 | Left Inferior Temporal Gyrus |
