## Supplementary Figure 3 for "A Human Gait Circuit Derived from Brain Lesions and Deep Brain Stimulation"

The topography of our stroke gait circuit was similar after controlling for seven other post-stroke deficits including motoricity, attention, right motor function, left motor function, spatial memory, verbal memory, and language.

Stroke gait circuit
(controlling for NIHSS)

Stroke gait circuit controlling
for average motoricity
spatial r = 0.88

Stroke gait circuit controlling
for attention
spatial r = 0.88

Stroke gait circuit controlling
for R motor function
spatial r = 0.67

Stroke gait circuit controlling
for L motor function
spatial r = 0.87

Stroke gait circuit controlling
for spatial memory
spatial r = 0.81

Stroke gait circuit controlling
for verbal memory
spatial r = 0.70

Stroke gait circuit controlling
for language
spatial r = 0.72

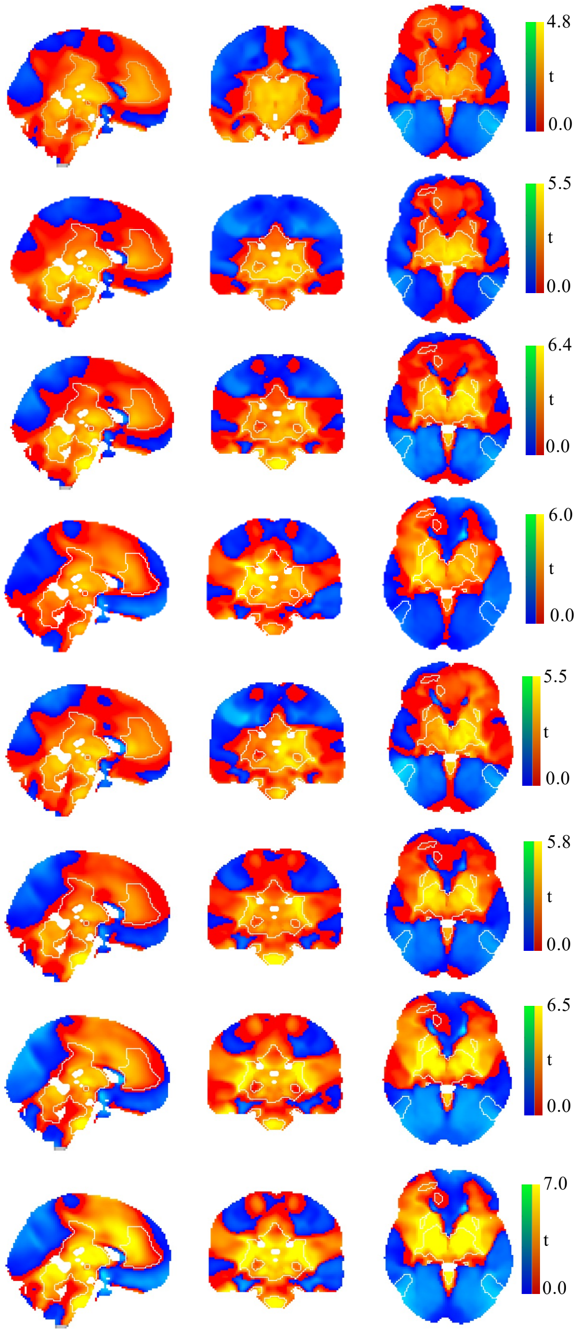
