## Supplementary Figure 4 for "A Human Gait Circuit Derived from Brain Lesions and Deep Brain Stimulation"

Data-driven circuits were generated for seven other cognitive and motor variables. Data-driven circuits for these seven post-stroke deficits failed to match the topography of our stroke gait circuit, with attention being the most similar (spatial r = -0.50).

Stroke Gait Circuit
(controlling for NIHSS)

Motoricity Circuit
spatial r = 0.32

Attention Circuit
spatial r = -0.50

R Motor Function Circuit
spatial r = -0.49

L Motor Function Circuit
spatial r = -0.08

Spatial Memory Circuit
spatial r = -0.26

Verbal Memory Circuit
spatial r = -0.19

Language Circuit
spatial r = 0.15

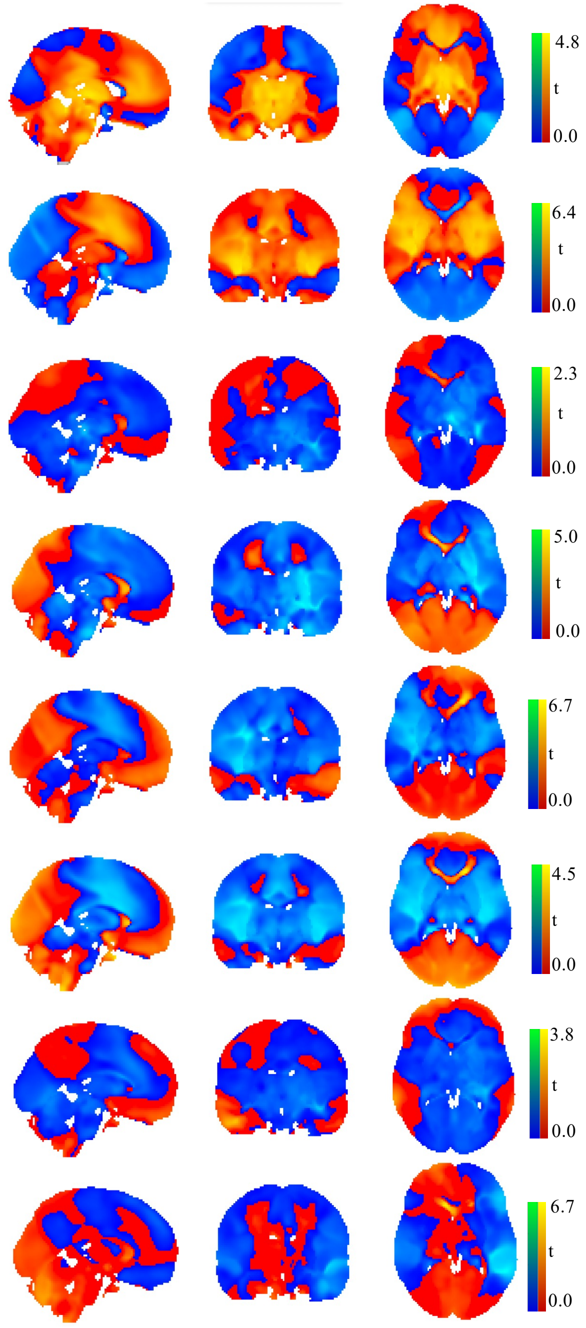
