## Supplementary Figure 5 for "A Human Gait Circuit Derived from Brain Lesions and Deep Brain Stimulation"

The topography of our DBS gait circuit was similar after controlling for three other motor features including lower extremity bradykinesia, lower extremity rigidity, and postural instability.

LE = lower extremity

DBS Gait Circuit
(controlling for UPDRS)

DBS Gait Circuit Controlling
for LE Bradykinesia
spatial r = 0.96

DBS Gait Circuit Controlling
for LE Rigidity
spatial r = 0.97

DBS Gait Circuit Controlling
for Postural Instability
spatial r = 0.96

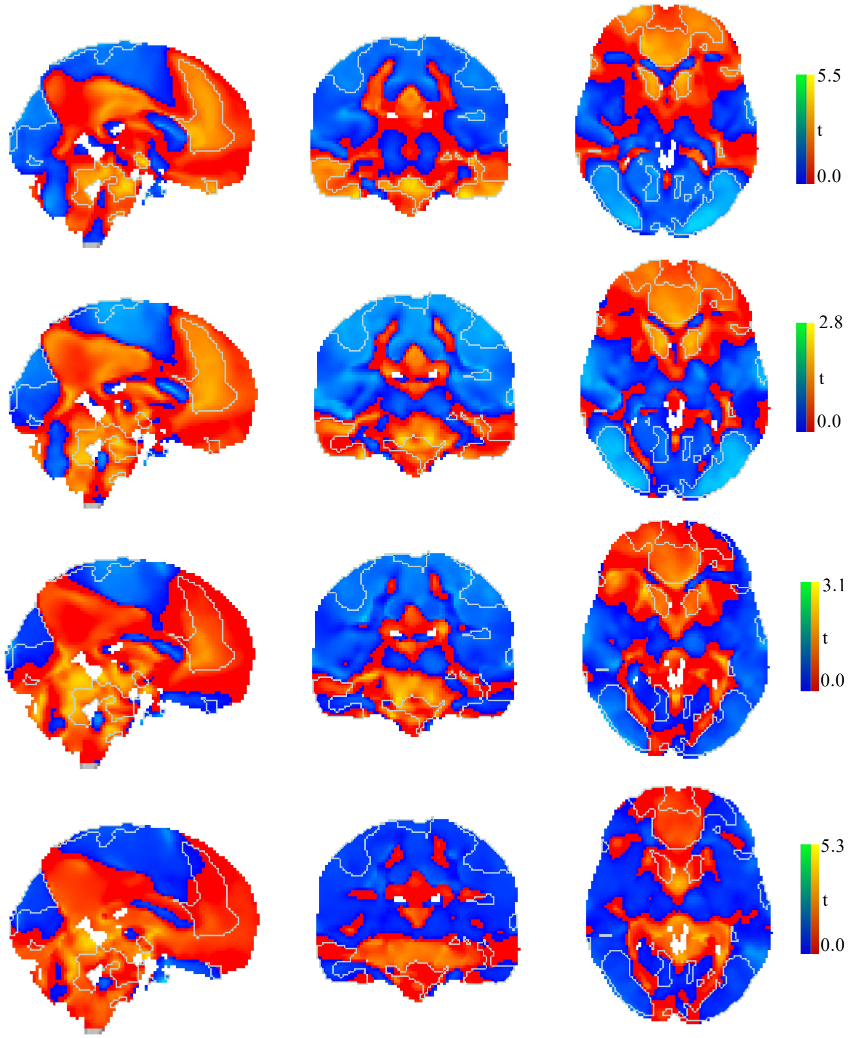
