## Supplementary Figure 6 for "A Human Gait Circuit Derived from Brain Lesions and Deep Brain Stimulation"

Data-driven brain circuits were generated for lower extremity bradykinesia, lower extremity rigidity, and postural instability. Lower extremity bradykinesia contributed the most to our DBS gait circuit as it shares a high spatial topography (spatial r = 0.90). LE = lower extremity

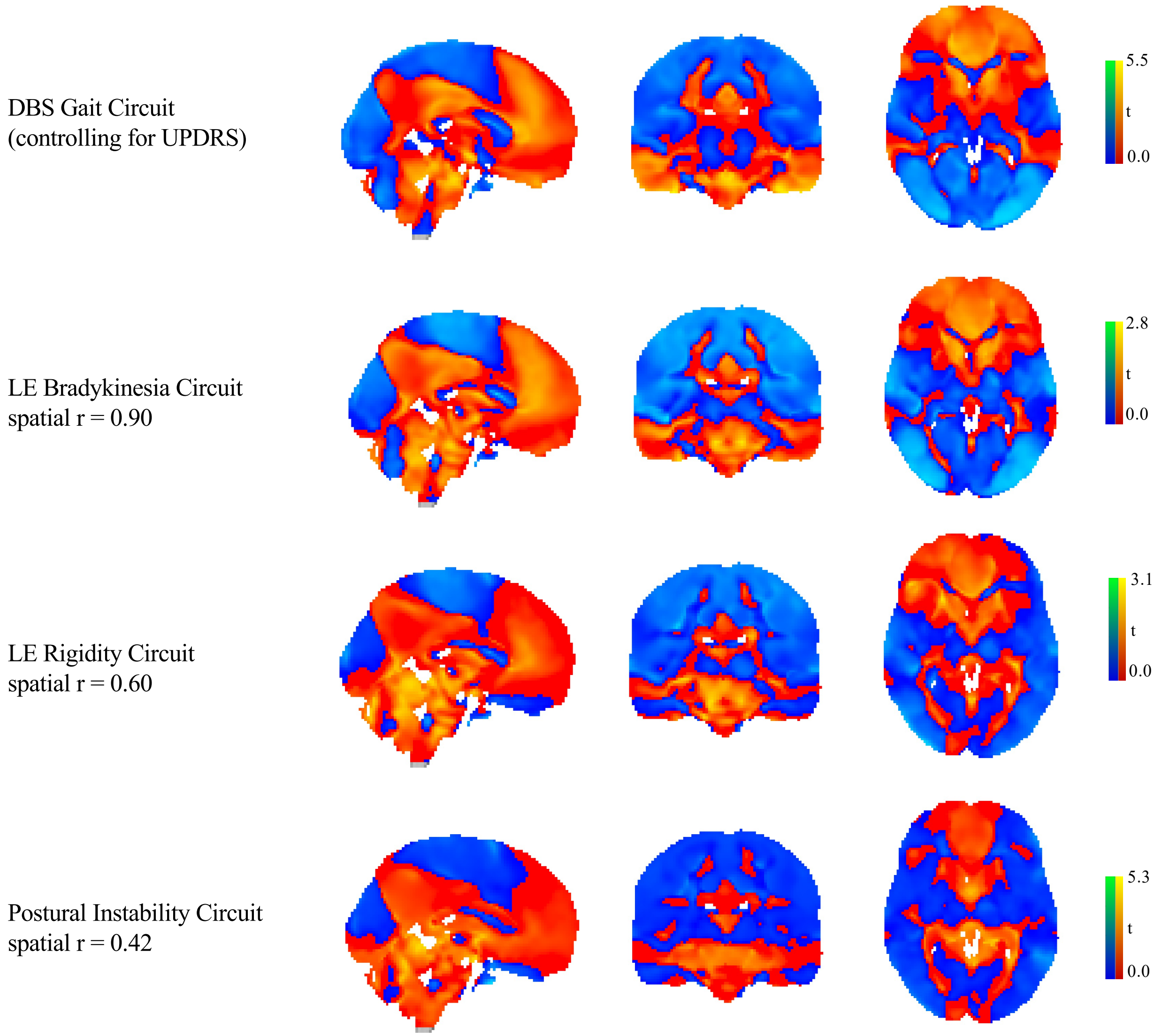
